# Long-Term Persistence of IgG Antibodies in recovered COVID-19 individuals at 18 months and the impact of two-dose BNT162b2 (Pfizer-BioNTech) mRNA vaccination on the antibody response

**DOI:** 10.1101/2022.01.18.22269349

**Authors:** Puya-Dehgani-Mobaraki, Chao Wang, Alessandro Floridi, Emanuela Floridi, Asiya K Zaidi

**Affiliations:** Associazione Naso Sano, Umbria Regional Registry of volunteer activities, Italy Via Luca Benincasa 2, San Mariano, 06073, Perugia-Italy; Faculty of Health, Social Care and Education, Kingston University and St George’s, University of London, London, SW17 0RE, UK; Laboratory of Nuclear Lipid BioPathology, Centro Ricerche Analisi Biochimico Specialistiche, Perugia, Italy; Associazione Naso Sano, Italy

## Abstract

This era of emerging variants needs a thorough evaluation of data on the long-term efficacy of immune responses in vaccinated as well as recovered individuals, to understand the overall evolution of the pandemic. In this study, we aimed to assess the dynamics of IgG titers over 18 months in n=36 patients from the Umbria region in Italy, who had a documented history of COVID-19 infection in March 2020, and then compared the impact of two-dose BNT162b2 (Pfizer-BioNTech) vaccination on the antibody titers of these patients with the ones who did not receive any dose of vaccine. This is the longest observation (March 2020-September 2021) for the presence of antibodies against SARS-CoV-2 in recovered individuals along with the impact of 2 dose-BNT162b2 vaccination on the titers. Fixed-effect regression models were used for statistical analysis which could be also used to predict future titer trends. At 18 months, 97% participants tested positive for anti-NCP hinting towards the persistence of infection-induced immunity even for the vaccinated individuals. Our study findings demonstrate that while double dose vaccination boosted the IgG titers in recovered individuals 161 times, this “boost” was relatively short-lived. The unvaccinated recovered individuals, in contrast, continued to show a steady decline but detectable antibody levels. Further studies are required to re-evaluate the timing and dose regimen of vaccines for an adequate immune response in recovered individuals.

## Introduction

This era of emerging variants needs a thorough evaluation of data on the long-term efficacy of immune responses in vaccinated as well as recovered individuals, to understand the overall pandemic evolution. Vaccine equity is an evolving global challenge that needs prompt alternate strategies for equal allocation across all countries in line with WHO’s campaign for Vaccine Equity. [1] Recent data has suggested post-infection immunity to be durable, long-acting, and protective. [2-8]. However, recovered individuals have also been advised to receive complete vaccination similar to naïve individuals with no previous history of COVID-19 infection. Whether two doses are required in recovered individuals or a single dose could offer adequate protection, is still a matter of debate.

Real-time data indicates that recovered individuals have protective immunity that could be sufficiently enhanced by a single dose of the vaccine rather than a double dose. Interestingly, neither recall responses nor ideal vaccine dosing regimens have been studied in them. [9,10] When compared to natural infection, vaccination elicits a more specific response which is greater in magnitude and is largely focused on the spike-receptor binding domain (S-RBD) rather than Nucleocapsid (NCP). Recent data suggests impaired protection, predominantly viral neutralization, and vaccine effectiveness against the variants of concern. [11]

In this study, we aimed to assess the dynamics of IgG titers over 18 months in patients who had a documented history of COVID-19 infection in March 2020, and then compared the impact of two-dose BNT162b2 (Pfizer-BioNTech) vaccination on the antibody titers of these patients with the ones who did not receive any dose of vaccine (NCT05038475). [12]

## Materials and methods

### Patient cohort

A monocentric longitudinal observational study was conducted in patients based in the Umbria region of Italy who had tested positive for SARS-CoV-2 in March 2020 by RT-qPCR, aimed to analyze the presence of antibodies against SARS-CoV-2. These RT-PCR tests were performed by the Local health regulatory authorities according to the national guidelines and standard operating protocols. The patients were managed as per the set protocols by the treating doctor, based on the severity of the disease. On recovery, all subjects were informed about this study for seroprevalence and were invited for voluntary participation. After detailed written informed consent, serological samples were collected at timed intervals and antibody titers were analyzed using the MAGLUMI® 2019-nCoV lgM/lgG chemiluminescent analytical system (CLIA) assay and the MAGLUMI® SARS-CoV-2 S-RBD IgG CLIA. (New Industries Biomedical Engineering Co., Ltd [Snibe], Shenzhen, China). Both these immunoassays were granted Emergency Use Authorization by the US Food and Drug Administration, anti-NCP during the initial months of the pandemic followed by anti-S-RBD later in 2020. [13]

Using a standardized questionnaire, the participants were asked to provide information about their COVID-19 clinical history along with symptoms and treatment undertaken. Although reminders were sent for follow-up periodically, participation in the study was completely voluntary. The study participants did not receive any compensation or any other benefit but were informed individually about their antibody status.

The blood samples were collected after informed consent by the patients and with the approval of the ethics committee of the Associazione Naso Sano (Document number ANS-2020/001) at an accredited lab (Laboratory of Nuclear Lipid BioPathology, CRABION, Perugia, Italy). Data collection and analysis were masked from the main principal investigator, who was also a part of the study sample to avoid observer bias. [14,15] The study was conducted in accordance with the Declaration of Helsinki and national and institutional standards.

## Patient selection

Study timeline and assays used: The antibody titers were analyzed in two phases, based on the availability of immunoassays at the time, i.e from May 2020 to January 2021, antibodies against NCP were analyzed using the MAGLUMI® 2019-nCoV lgM/lgG CLIA assay through sequential serum samples. Time was treated as a factor and six different time points were defined; (T0-T5). The first blood sample was collected in May 2020, 2 months after infection (march), and was defined as T0. Consecutive serological samples were analyzed at three months (T1), five months (T2), seven months (T3), eight months (T4) ten months (T5) post-infection in June, August, October, November of 2020, and January 2021 respectively.

The second phase comprised of the introduction of a different immunoassay; MAGLUMI® SARS-CoV-2 S-RBD IgG CLIA to detect anti-S-RBD antibodies from Feb 2021 to September 2021.

Analysis using both the immunoassays (anti-NCP and anti-S-RBD) for each patient of the study group could not be done throughout 18 months due to practical challenges associated with it. From the original cohort, in n=24 individuals, anti-NCP was analyzed from May 2020 to January 2021 and then anti-S-RBD was analyzed from February 2021 to September 2021. For n=12 (8 female and 4 male) participants, who met the eligibility criteria for participation (documented history of COVID-19 in March 2020) that were enrolled in this study in Feb 2021, only anti-S-RBD could be analyzed from February 2021 to September 2021 making the final sample size of n=36 participants.

## Introduction of the vaccinated subgroup

Since the legal provisions adopted by the Italian Ministry of Health advised vaccination, irrespective of previous disease status, n=21 participants were gradually vaccinated from mid-March 2021. Out of these, n=19 participants continued to follow up voluntarily for antibody titer assessment at monthly intervals.

Therefore at 18 months, the study participants were divided into two groups:

*Group A:* Recovered and received two doses of BNT162b2 vaccine (n=21). From this recovered vaccinated group, n=2 (P23 and P30 in the figure) failed to follow up post-vaccination making the final sample size of n=19.

*Group B:* Recovered individuals who were unvaccinated (n=15).

For the month of September 2021 (18 months post-infection), the immunoassay to detect anti-NCP was reintroduced so that even vaccinated individuals could be analyzed for the persistence of infection-induced immunity not generated by mRNA spike-based vaccination. Only qualitative assessment could be done for the presence of anti-NCP antibodies at this point.

Although the study sample is small, the presented data can be useful in providing information on the longer-term dynamics of relevant antibodies since the beginning of the pandemic and the impact of vaccination on antibody responses.

## Analytical system used

For analysis at 18 months, the study participants were divided into two groups based on their vaccination status.

Group A: recovered and received two doses of BNT162b2 vaccine (n=19) and Group B: recovered individuals who were unvaccinated (n=15).

As per the Specifications, the anti-SARS-CoV-2 S-RBD IgG assay had a sensitivity of 100% with CI [99.9%-100.0%] at ≥15 days post symptom onset and specificity of 99.6%; CI [98.7%-100.0%].

High concentration samples were diluted automatically by analyzers and the recommended dilution was 1:9 with the diluent in the kit. The sample, buffer, and magnetic microbeads coated with S-RBD recombinant antigen were mixed thoroughly and incubated, forming immune complexes. After precipitation, decanting of supernatant, and performing a wash cycle, ABEI labeled with anti-human IgG antibody was added, and incubated to form complexes. Again after precipitation in a magnetic field, decanting of supernatant, and performing another wash cycle, the Starter 1+2 were added to initiate a chemiluminescent reaction. The light signal was measured by a photomultiplier as relative light units (RLUs), which is proportional to the concentration of SARS-CoV-2 S-RBD IgG presented in the sample. The measurements and interpretation of results were made according to the manufacturer’s instructions. The analyzer automatically calculates the concentration in each sample using a calibration curve which is generated by a 2-point calibration master curve procedure. The results were expressed in AU/mL. A result less than 1.00 AU/mL (<1.00 AU/mL) was considered to be non-reactive while a result greater than or equal to 1.00 AU/mL (≥1.00 AU/mL) was considered to be reactive. [16,17]

## Statistical Analysis

To model how antibody response evolves over time, we used a fixed-effects linear regression while adjusting for the testing method/immunoassay used (NCP or S-RBD), vaccination status, and temporal effects. To model the non-linear effects of time, a restricted cubic spline function for time (month) was applied in the regression model using the method by Orsini and Greenland (2011) to visualize and interpret the results. [18] To account for serial correlation, the error term is modeled as a first-order autoregression (AR) process.

## Results

Figure 1 shows how the IgM and IgG titers have evolved over time for the recovered subjects and the impact of two-dose BNT162b2 (Pfizer-BioNTech) vaccination on the antibody titers.

**Figure 1.**
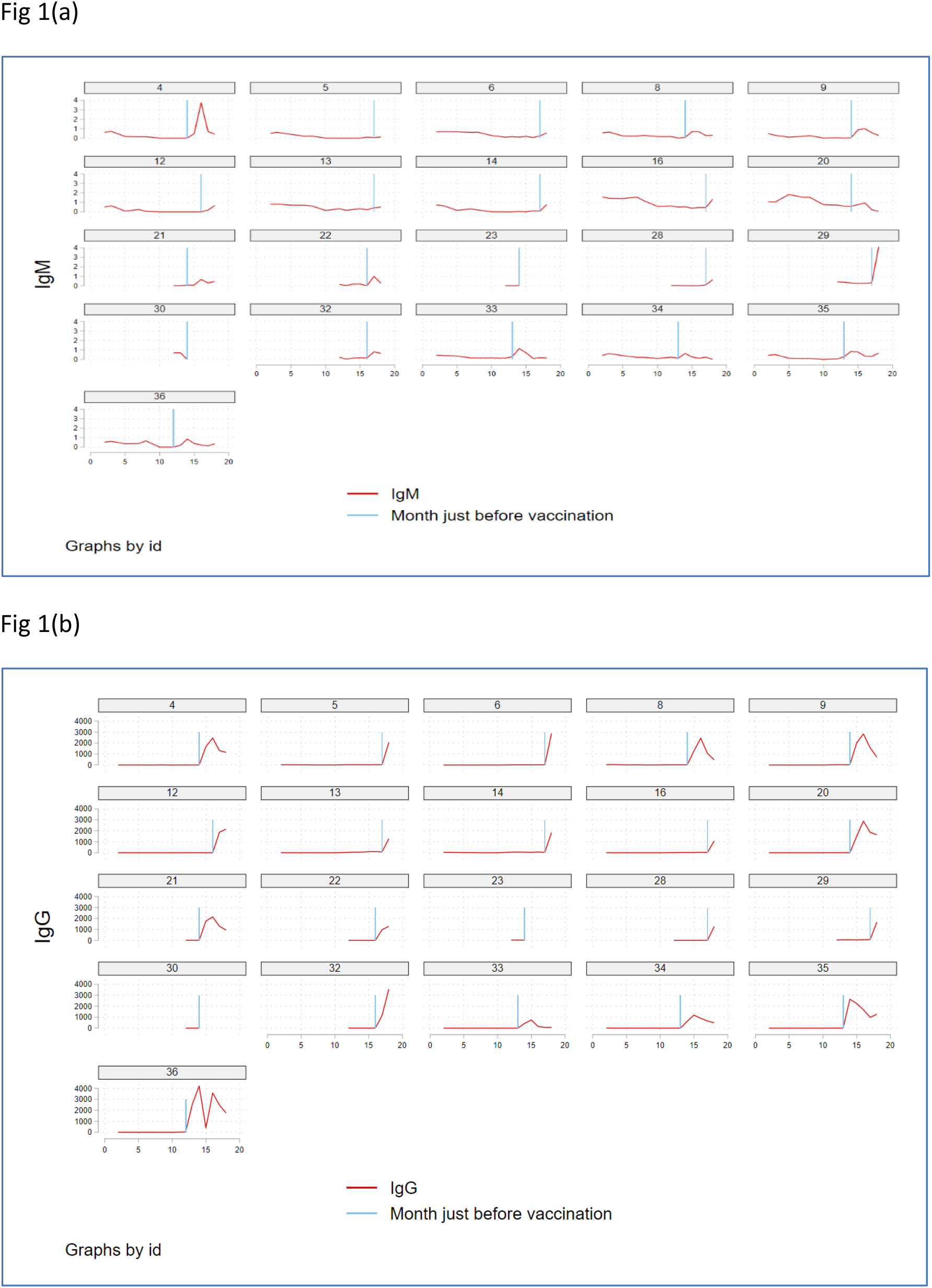
Figure 1 shows how the IgM and IgG titers have evolved over 18 months for n=21 of Group A : recovered subjects and the impact of two-dose BNT162b2 (Pfizer-BioNTech) vaccination on the antibody titers, where Figure 1(a) shows the IgM titer trend (red) and Figure 1(b) shows the IgG titer trend (red line) over 18 months in each subject as per the patient number that was assigned randomly. The blue vertical line represents the time of vaccination. From this recovered vaccinated group, n=2 (P23 and P30 in the figure) failed to follow up post-vaccination.

Figure 1(a) shows the IgM titer trend (red) over 18 months in each subject as per the patient number that was assigned randomly. The blue vertical line represents the time of vaccination. Figure 1(b) shows the IgG titer trend (red) over 18 months in each subject as per the patient number that was assigned randomly. The blue vertical line represents the time of vaccination. The graphs demonstrate that vaccination has an enormous impact on antibody titers. It was interesting to note that, although there was a rapid and steep rise in the antibody titers immediately post-vaccination, this rise was short-lived. This explains the role of boosters that might, perhaps “boost” the titer levels for a brief duration. Whether the titer trend after vaccination drops back to pre-vaccination levels in a few weeks or in fact, lowers below the baseline needs further analysis on a larger population.

Fig 2(a) visualizes the modeling results for the four scenarios, based on immunoassay employed (anti-NCP or anti-S-RBD) and completion of two doses of BNT162b2 (Pfizer–BioNTech) mRNA vaccine: 1) unvaccinated and tested against NCP; 2) vaccinated and tested against NCP; 3) unvaccinated and tested against S-RBD; and 4) vaccinated and tested against S-RBD. IgG titers were log-transformed to avoid negative values.

**Figure 2.**
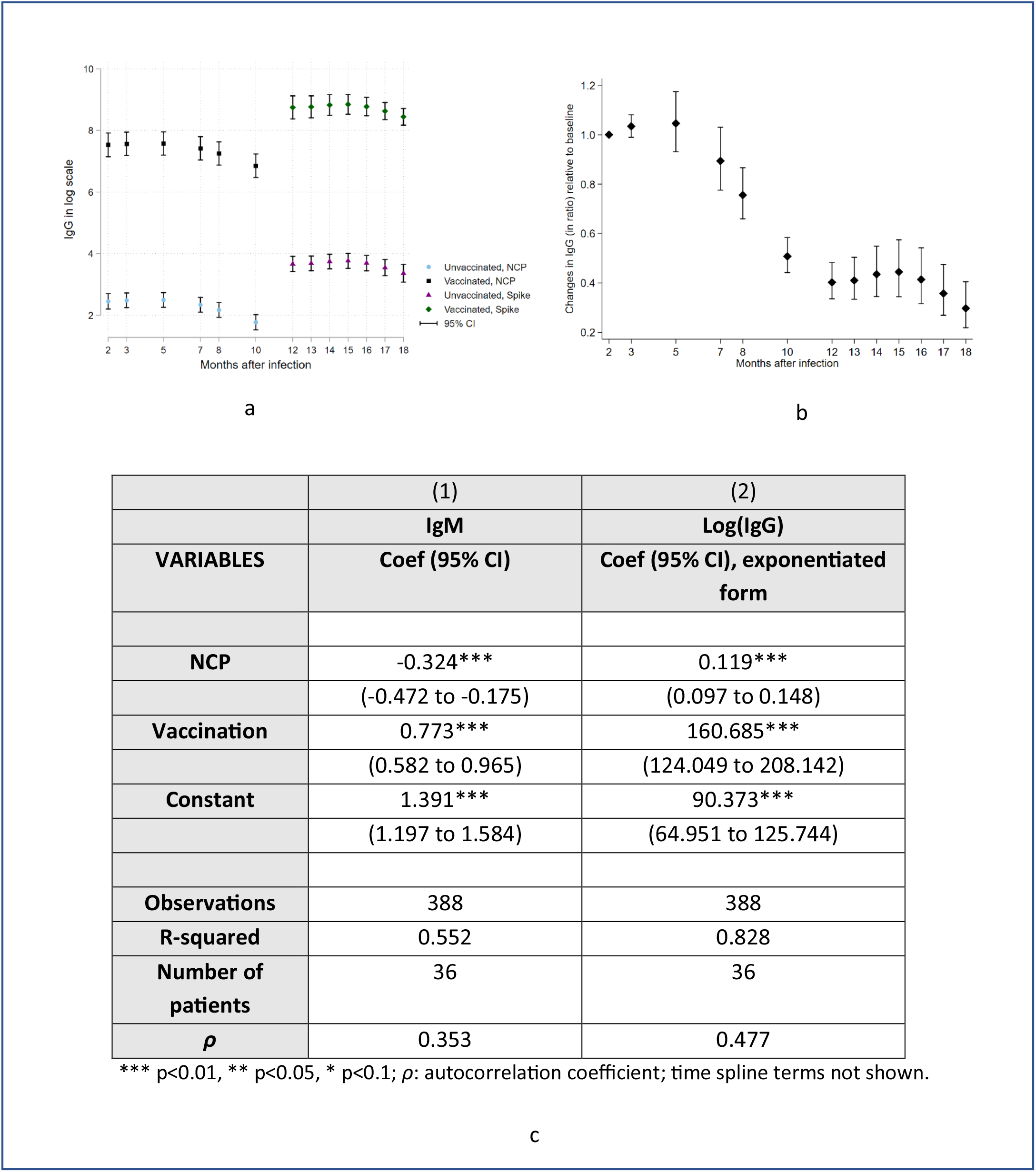
Fig 2(a) visualizes the modeling results for the four scenarios, based on immunoassay employed (anti-NCP or anti-S-RBD) and completion of two doses of BNT162b2 (Pfizer–BioNTech) mRNA vaccine: 1) unvaccinated and tested against NCP; 2) vaccinated and tested against NCP; 3) unvaccinated and tested against S-RBD; and 4) vaccinated and tested against S-RBD. IgG titers were log-transformed to avoid negative values. Fig 2(b) describes the differences in antibody responses between a given month and the baseline month (month 2), holding all the other variables (vaccination status and testing method) constant and Fig 2(c) describes the modeling results to determine the effect of vaccination on antibody titers (IgM and IgG) in recovered subjects. NCP has lower IgM, by amount of 0.324, relative to spike. Similarly, NCP has a lower IgG than spike by a factor of 0.119. The vaccines lead to large increase in antibody responses: vaccination increases IgM by amount of 0.773, and IgG by 161 times.

Fig 2(b) describes the differences in antibody responses between a given month and the baseline month (month 2), holding all the other variables (vaccination status and testing method) constant. This allows visualization of antibody response dynamics regardless of other factors since this model assumes an independent temporal effect. Statistical models (especially *Fixed-effect* regression models) are flexible to *diminish* the *impact* of confounding by (unmeasured) fixed/time-invariant factors.

Fig 2(c) describes the modeling results to determine the effect of vaccination on antibody titers (IgM and IgG) in recovered subjects. NCP has lower IgM, by amount of 0.324, relative to spike. Similarly, NCP has a lower IgG than spike by a factor of 0.119.

A double-dose vaccination with BNT162b2 in recovered individuals induced an enormous but short-lived increase in antibody titers. While the IgM titers increased by an amount of 0.773, the IgG titers increased 161 times.

However, we do agree that a caveat should be noted that the model assumed that anti-S-RBD titers only affect the level but not the trend/shape of the curve relative to anti-NCP. Further analysis of the interaction between time and type of antibody is not statistically significant which supports our original model.

It was interesting to note that all participants from Group B (n=15) tested positive for anti-S-RBD for 18 months post-infection, with no case reported of reinfection despite multiple waves of mutant strains encountered in the region since March 2020. Also, 33 out of the 34 participants (97%) tested positive for anti-NCP at 18 months. We hypothesize that anti-NCP immunoassays could be a valuable tool for the assessment of persisting infection-induced immunity and should be re-adopted for analysis even for vaccinated individuals. In terms of strain, whether the infection with the wild-type conferred a more robust protective immunity against re-infection as compared to infection with other mutant strains needs further large-scale analysis.

## Discussion

In this study, we longitudinally analyzed the antibody responses in patients that recovered from COVID -19, 18 months after SARS-CoV-2 infection (wild type), and divided them into two groups for comparison based on vaccination with two doses of BNT162b2. To our knowledge, this is the longest observational study that reports the persistency of antibodies against SARS-CoV-2 at 18 months and the impact of 2 dose BNT162b2 vaccine in seropositive convalescent patients.

Our observations have been in line with previously published studies suggesting the presence and persistence of long-term protective immunity after SARS-CoV-2 infection. A study by Gaebler et al concluded that “memory B cell response to SARS-CoV-2 evolves between 1.3 and 6.2 months after infection in a manner that is consistent with antigen persistence”. [2] In another study by Abu-Raddad LJ et al (Qatar Study), it was observed that the “estimated efficacy of natural infection against reinfection was 95% over 7 months”. [3] Anand et al. also concluded that “anti-Spike and anti-receptor binding domain (RBD) immunoglobulin M (IgM) in plasma decay rapidly, whereas the reduction of IgG is less prominent up to 8 months”. [4] A study conducted on 3015 healthcare workers by Egbert ER. et al demonstrated “durability of spike antibodies to SARS-CoV-2 up to 10 months after natural infection”. [5] Another interesting study by Turner et. al (n=77, mild infection) concluded “anti-S-IgG to be detectable at 11 months; S-binding bone marrow plasma cells (BMPCs) are quiescent, indicating that they are part of a long-lived compartment”. [6] Similarly, Gallais et al. observed “a long-term persistence of anti-RBD antibodies that may reduce risk of reinfection up to 13 months. [7] Our previous study for infection-acquired immunity against SARS-CoV-2 at 14 months has been published. [8]

Vaccination-induced immunity in individuals with no history of previous infection has been relatively short-lived (half-life of 69-173 days) with requirements of boosters to augment antibody titers against subsequent variants of concern, especially the Omicron variant. [19] This variant harbors more mutations compared to prior variants, and therefore “efficiently” escapes humoral immunity induced by primary vaccination. It was observed by Garcia-Beltran et al that additional mRNA vaccine doses (boosters) tend to demonstrate cross-neutralizing responses against Omicron as well. This could be due to “affinity maturation of the existing antibodies” or new shared epitopes of variants serving as targets. [20] The authors also observed a 9-fold increase in the geometric mean neutralization titer (GMNT) for Delta pseudoviruses in individuals that received three-doses of BNT162b as compared to two doses, and an increase in cross-neutralization of the Omicron variant by 27-fold for individuals who received three doses of BNT162b vaccine.

This neutralization signal could be correlated with anti-SARS-CoV-2 spike antibody titers measured by anti-spike immunoassays that are widely available. [21] Therefore, our focus on anti-spike antibody titers is valuable.

Our study findings demonstrate that while double dose vaccination boosted the IgG titers in recovered individuals 161 times, this “boost” was relatively short-lived. The unvaccinated recovered individuals, in contrast, continued to show a steady decline but detectable antibody levels.

Whether or not the present vaccines induce sterilizing immunity remains unclear. They trigger serological IgA response but do not generate mucosal IgA at the virus site of entrance. Vaccine breakthrough infections may be because of deficient local preventive immunity due to absence of mucosal IgA. [22]

Upon mRNA BNT162b2 vaccination, neutralizing antibodies are boosted significantly in the serum, but not at the mucosal sites and saliva. This indicates poor activation of oral mucosal immunity that fails to limit the viral attachment to target cells at the entry site. [23]

In contrast, convalescent individuals that recovered from COVID-19 infection (also known as hybrid immunity) demonstrated the presence of specific IgA indicating that natural infection conferred mucosal immunity. [22,23 24]

Vaccination-induced immunity in individuals with a history of prior infection was associated with high levels of wild-type neutralization titers even when vaccinated distantly. (>6 months)[25]

In line with a study by Hall et al, our study results demonstrate that a previous COVID-19 infection (infection acquired immunity) appeared to elicit robust, long-term, and sustained levels of SARS-CoV-2 antibodies in individuals who received two doses of BNT162b2. It was also observed that individuals with hybrid immunity developed the “highest and most durable protection”. [26]

Our study had some limitations. First, a small sample size. Second, ideally, simultaneous antibody titer detection of each patient at each time point using both NCP and S-RBD assay would have given the best results for comparative analysis but the S-RBD assay received emergency approval only later in 2020.

The nucleocapsid protein (NCP) of the SARS-CoV-2 is considered a biomarker associated with natural exposure. Since this NCP is not present in spike-based vaccines, there is no vaccine-induced response against it in vaccinated individuals. Testing for anti-NCP could be an important tool to analyze the type of existing immunity in an individual (vaccine-induced/ natural/ hybrid). Moreover, Vaccinated individuals who were previously exposed usually test positive for antibodies against NCP (hybrid immunity) while the vaccinated individuals who have never been exposed lack anti-NCP antibodies (vaccine-induced immunity). [27,28]

The major strengths of our study include the longest observation (March 2020-September 2021) for the presence of antibodies against SARS-CoV-2 in recovered individuals along with the impact of 2 dose-BNT162b2 vaccination on the titers. Secondly, fixed-effect regression models were used for statistical analysis that diminishes the impact of confounding by (unmeasured) fixed/time-invariant factors. These modeling results could also be used to predict future titer trends. Thirdly, since the immunoassay to detect anti-NCP was reintroduced at 18 months, even the vaccinated individuals could be analyzed for the persistence of infection-induced immunity not generated by mRNA spike-based vaccination. Finally, combining the data for both was necessary because this makes it possible to explore the differences between anti-NCP and anti-S-RBD (as also shown by the regression results in Fig 2c). This is similar to an interrupted time series (ITS) analysis which typically looks at how an ‘intervention’ affects the temporal pattern between before and after periods using whole datasets. Our data is rich in that it is both cross-sectional (multiple patients) and time-series which allows us to adopt complex longitudinal models.

## Conclusion

Further studies are required to re-evaluate the timing and dose regimen of vaccines for an adequate immune response in recovered individuals.

## Data Availability

All data produced in the present study are available upon reasonable request to the authors

## Acknowledgment

The authors would like to acknowledge the assistance provided by the Rotary Club Fortebraccio Montone for the successful conduction of this research study.

